# Medical Costs of Keeping the US Economy Open During COVID-19

**DOI:** 10.1101/2020.07.17.20156232

**Authors:** Jiangzhuo Chen, Anil Vullikanti, Stefan Hoops, Henning Mortveit, Bryan Lewis, Srinivasan Venkatramanan, Wen You, Stephen Eubank, Madhav Marathe, Chris Barrett, Achla Marathe

## Abstract

We use an individual based model and national level epidemic simulations to estimate the medical costs of keeping the US economy open during COVID-19 pandemic under different counterfactual scenarios. We model an unmitigated scenario and 12 mitigation scenarios which differ in compliance behavior to social distancing strategies and to the duration of the stay-home order. Under each scenario we estimate the number of people who are likely to get infected and require medical attention, hospitalization, and ventilators. Given the per capita medical cost for each of these health states, we compute the total medical costs for each scenario and show the tradeoffs between deaths, costs, infections, compliance and the duration of stay-home order. We also consider the hospital bed capacity of each Hospital Referral Region (HRR) in the US to estimate the deficit in beds each HRR will likely encounter given the demand for hospital beds. We consider a case where HRRs share hospital beds among the neighboring HRRs during a surge in demand beyond the available beds and the impact it has in controlling additional deaths.

## 1 Introduction

As states push to end social distancing and reopen businesses, it is important to understand the cost of opening in terms of lives lost and medical costs incurred. A premature opening will likely cause more deaths and infections as the healthcare system will likely get overwhelmed, and may wipe out all the gains made in the first shutdown. We use an agent-based model and simulation framework to estimate the immediate medical cost of COVID-19 under different mitigation scenarios. The scenarios consider social distancing with different durations and varying compliance levels. The simulation framework uses a detailed representation of the US population and their social interactions to study the spread of COVID-19. An SEIR (susceptible-exposed-infected-recovered) model captures the time varying health states of the individuals. The infected individuals arrive at one of the three health states i.e. medically attended, hospitalized, or ventilated before getting to the final health state i.e. recovered or dead, as shown in figure 1.

**Figure 1.**
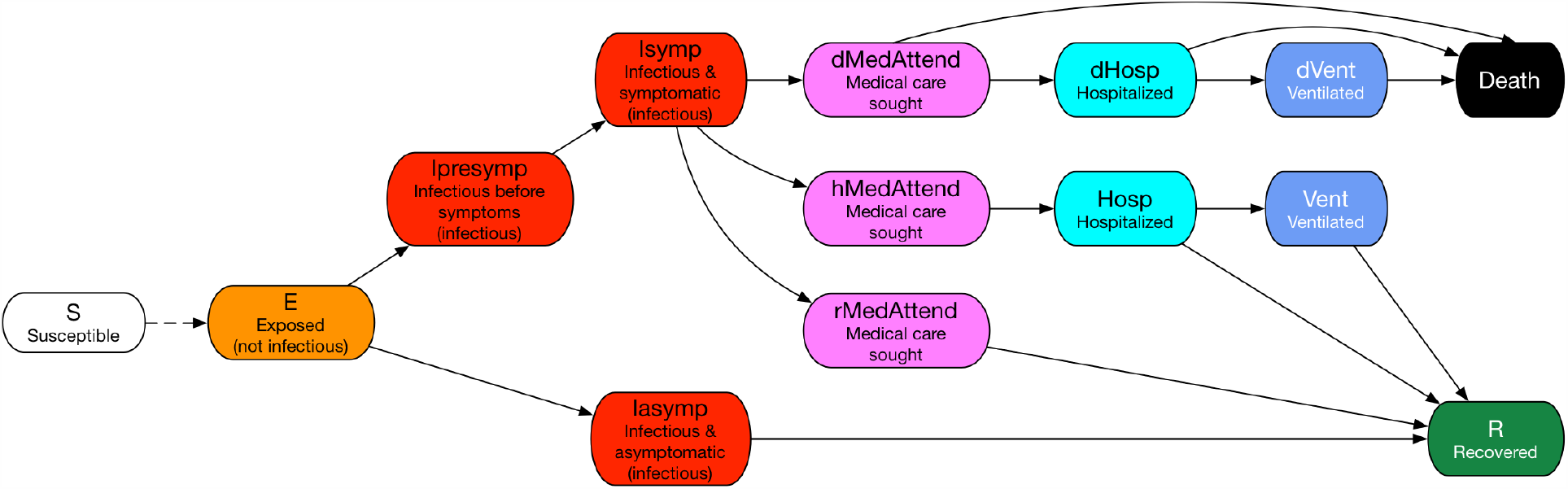
State transitions in the COVID-19 disease model.

Medical costs are applied based on the three health states i.e. medically-attended, hospitalized and ventilated. In addition, if an infected individual dies, then the “value of statistical life” is used to estimate the cost of death. We also estimate the shortage of hospital beds that is likely to occur in each Hospital Referral Region (HRR) given the demand for hospital beds and the number of available beds in each HRR in the US. Data on the number of beds in each HRR is obtained from the American Hospital Association (AHA) and a fraction of them are assumed to be available for COVID-19 patients. We consider cases where neighboring HRRs share or do not share hospital beds during a surge in demand.

This information is then used to calculate additional deaths and medical costs for each of the mitigation scenarios. Policy makers can apply this kind of analysis to decide where the temporary hospitals may need to be built to offset the deficit in demand for beds. Our goal is to use this knowledge to provide guidance to public health officials and policy makers on the trade-offs between the length of lockdown, compliance to social distancing, infections, deaths and the medical costs. Our scenario-based analysis estimates the burden of the disease in monetary terms, and helps rank-order mitigation strategies.

In related work, authors in^1^ consider potential health care costs and resource usage under different attack rates which vary from 20% to 80%. However, it does not consider any interventions or mitigation strategies. Our research focuses on counterfactual mitigation scenarios and their respective costs. We use recent cost estimates for COVID-19 available from the Kaiser Family Foundation^2^ which uses cost of pneumonia cases as a proxy. Our detailed network based model, that captures heterogeneous social interactions and contact times among the individuals in the population, is one of the unique features of the analysis. Additionally, no other research has provided an estimation of medical costs for such detailed mitigation scenarios for the entire US.

Our results show that (i) Without mitigation, the total medical costs would be a significant fraction (5%) of the US GDP; (ii) a lockdown of just two months, if done early in the epidemic, and with sufficient compliance, could have reduced the medical costs by more than 90%; (iii) if 90% compliance could be achieved, then even a 45 day lockdown period would have been enough to contain the epidemic and the medical costs; (iv) if HRRs do not share hospital beds with other HRRs, a significant deficit of beds will cause medical costs to skyrocket, through increase in deaths. However, if HRRs shared beds with their neighboring HRRs, the bed-deficits and additional deaths could be reduced to almost zero; and (v) a sensitivity analysis of the parameters shows the costs are most sensitive to the duration of the stay-home order.

## 2 Data and Methodology

We build on our modeling and simulation framework for epidemic spread^3–9^ using an individual level synthetic social contact network^5,10^—which represents each individual in the population along with their demographic attributes (e.g., age, gender, income), and their social interactions. The main steps in the first-principles based construction of synthetic populations and social contact networks are: (i) construct a synthetic population by using US Census and other commercial databases; (ii) assign daily activities to individuals within each household using activity and time-use surveys (American Time Use Survey data and National Household Travel Survey Data); (iii) assign a geo-location to each activity of each person based on data from Dun and BradStreet, land-use, Open Street Maps etc.; and (iv) construct a dynamic social co-location based social contact network that is induced when people simultaneously visit locations. These networks have been validated and used for numerous public health analyses before such as^3,5,11–16^. For details on the construction of social networks, see^11,12,17^.

The SEIR disease model and parameters used here for COVID-19 have been defined in the *best guess 2020-04-14 version* of “COVID-19 Pandemic Planning Scenarios” document prepared by the Centers for Disease Control and Prevention (CDC) SARS-CoV-2 Modeling Team^18^. The sequence of health-state transitions and possible paths are shown in figure 1. There are many possible health states and paths an individual can move through as it transitions from susceptible to its final health state. This model is age stratified for the following categories i.e. preschool (0-4 years), students (5-17) adults (18-49), older adults (50-64) and seniors (65+) and calibrated for each of the age groups separately. Details on the transition probabilities between health states for each age group and the length of the stay in each health state are shown in the table in the Appendix. Our models and simulation framework have been used in some of our ongoing SARS-CoV-2 response work for the Virginia Department of Health and the US Department of Defense^19^.

### 2.1 Medical Costs

To estimate the medical cost of treating COVID-19 patients, we use the average cost of treatment for pneumonia, paid among “large employer health insurance” plans, as a proxy^2^. The costs for each health state are shown in table 1. Note that each infected individual’s medical cost is counted only once. For example if a person is in ventilated state, after having gone through “medAttend” and “Hosp” state, costs are cumulative to the “vent” state.

**Table 1.** Average cost of medical care under different health states^2^.

| Health state | Average medical treatment costs per person |
| --- | --- |
| Medically-Attended | \$9,763 (cost of treating pneumonia without complications) |
| Hospitalization | \$13,767 (cost of treating pneumonia with complications or comorbidity) |
| Ventilator | \$61,168 (cost of treating pneumonia with ventilator) |
| Death | \$2M (value of statistical life) |

#### Value of Life

The total medical costs can be measured through multiple criteria in terms of number of deaths and treatment costs, or a single criterion in dollar terms by converting deaths into dollars using “value of a statistical life”. There are various ways in which the value of a statistical life has been measured. US federal government uses $10 million dollar per life lost regardless of a person’s age while others have used estimates in the range of $160k to $2.4 million based on metrics like average lifetime earnings of a college graduate, 9/11 victim compensation, wrongful death claims, insurance policy value etc.^20–22^. We use an estimate of $2 million dollar as the value of life to convert deaths into costs^23^.

### 2.2 Interventions

We consider a number of mitigation scenarios that comprise of various social distancing strategies, compliance levels and durations of stay-home order. The following social distancing strategies are used:

1. Voluntary home isolation (VHI): Symptomatic people choose to stay at home (non-home type contacts are disabled) for 14 days.
2. School closure (SC): Schools and colleges are closed (school type contacts are disabled).
3. Stay home (SH): People follow public health “stay-home” directive (non-home type contacts are disabled). School closure and stay-home interventions start on different days in different states as stated in^24,25^. Once closed, schools are assumed to remain closed until end of August after which they reopen. Other social distancing interventions stop at 30, 45 or 60 days from the start date of the intervention, depending on the SH duration. Note that this implies all mitigation efforts end by the end of summer 2020.

Durations of stay-home order vary from 0, 30, 45 to 60 days. Compliance to stay-home and voluntary home isolation vary from 60%, 70%, 80% and 90%. Table 2 shows a factorial design with 12 mitigation scenarios and an unmitigated case, resulting in a total of 13 cells experiment. For each cell, 25 replicates are run and their averages reported. Table 3 shows a complete list of variables and their parameter values.

**Table 2.** Factorial design of mitigation scenarios: VHI and SH refer to “voluntary home isolation” and “stay-home” order respectively. Schools are assumed to remain closed until the end of August 2020 after which they reopen. VHI and SH interventions expire after 30, 45 or 60 days, based on the scenario.

| SH duration<br>(in days) | VHI and SH compliance rates |  |  |  |
| --- | --- | --- | --- | --- |
|  | 60% | 70% | 80% | 90% |
| 0 | None (unmitigated) |  |  |  |
| 30 | VHI_60_SH_60_30 | VHI_70_SH_70_30 | VHI_80_SH_80_30 | VHI_90_SH_90_30 |
| 45 | VHI_60_SH_60_45 | VHI_70_SH_70_45 | VHI_80_SH_80_45 | VHI_90_SH_90_45 |
| 60 | VHI_60_SH_60_60 | VHI_70_SH_70_60 | VHI_80_SH_80_60 | VHI_90_SH_90_60 |

**Table 3.**
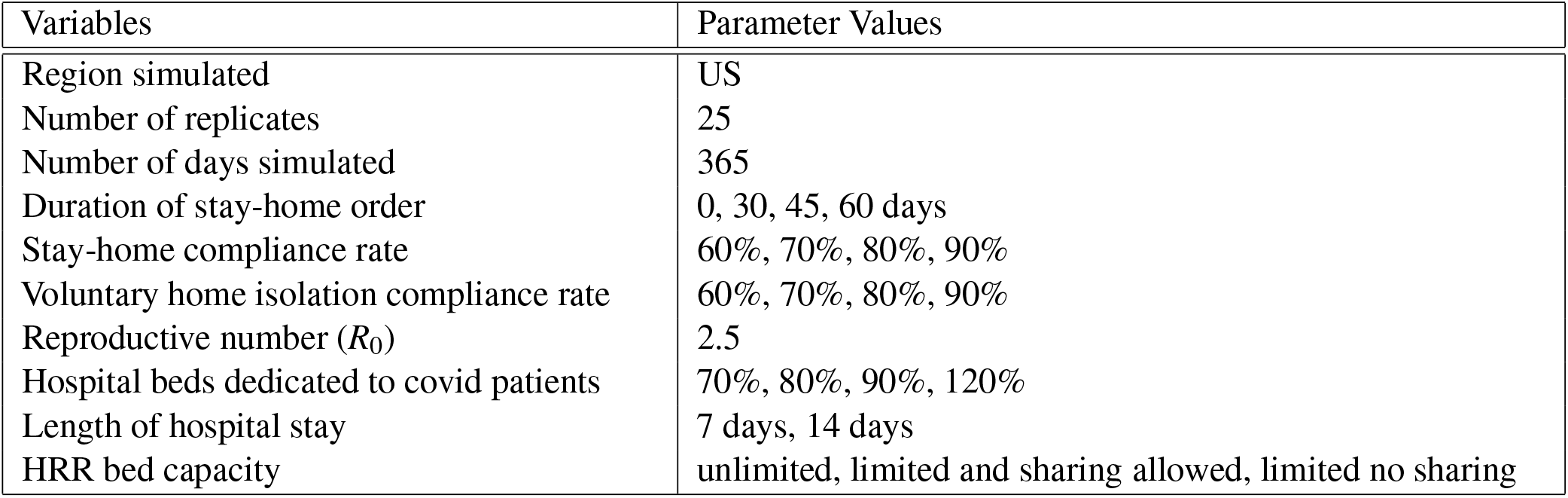
List of all the variables and their parameter values

| Variables | Parameter Values |
| --- | --- |
| Region simulated | US |
| Number of replicates | 25 |
| Number of days simulated | 365 |
| Duration of stay-home order | 0, 30, 45, 60 days |
| Stay-home compliance rate | 60%, 70%, 80%, 90% |
| Voluntary home isolation compliance rate | 60%, 70%, 80%, 90% |
| Reproductive number ($R_0$) | 2.5 |
| Hospital beds dedicated to covid patients | 70%, 80%, 90%, 120% |
| Length of hospital stay | 7 days, 14 days |
| HRR bed capacity | unlimited, limited and sharing allowed, limited no sharing |

### 2.3 Hospital Bed Capacity

We use hospital bed capacity data available for each HRR in the US from AHA, to calculate the deficits that are likely to be encountered by each region. We assume three scenarios regarding the use of hospital beds: (i) All COVID-19 patients who need a bed, will have one available i.e. there is no shortage of hospital beds, (ii) bed capacity in each HRR is limited and beds cannot be shared among hospitals in other HRRs to accomodate the surge in demand for beds and (iii) bed capacity in each HRR is limited but beds can be shared among hospitals in the neighboring HRR regions to accomodate the surge in demand.

Other factors considered in these scenarios are the average length of hospital stay of patients who are hospitalized, and the percentage of beds dedicated to COVID-19 patients in each HRR. Length of stay is 7 days or 14 days, and the percentage of beds available to COVID-19 patients is 70%, 80%, 90% or 120%. More than 100% bed capacity (120%) has been considered since many hospitals are able to temporarily increase their capacity beyond normal levels^26^.

Each day the demand for the number of beds in each HRR is determined by the simulations. The simulation results provide the counts of individuals who are in “hospitalized” state and depending upon the assumed duration of the hospital stay (7 days or 14 days), the demand for beds per day is calculated. Note that this includes individuals who are in “ventilated” state as well since everyone in “ventilated” state has to be in the “hospitalized” state first according to our disease model, see figure 1. The difference between the bed capacity and the counts of patients who need it, determines the deficit in hospital beds.

We assume that patients who need a hospital bed and cannot get it, will die. This is a strong assumption and hence four different values of dedicated bed capacities have been considered to show its sensitivity. Note that only the death counts and overall costs will change in scenarios (ii) and (iii) when limitations to bed capacity are considered. This assumption of “all bed deficits result in deaths” provides an upper bound on the medical costs resulting from the shortage of beds. If only a fraction of them die then the additional deaths and the costs can be appropriately scaled down.

## 3 Results

This section reports the simulation results and the medical costs that are likely to incur under various mitigation scenarios. For each of these scenarios we also consider four different possibilities for the availability of dedicated hospital beds for COVID-19 patients. Note that the hospital beds are considered as a post-processing step, after the simulations have been run and infected individuals have arrived at “hospitalized” state.

Figure 2 shows the epidemic curves with daily new infections in the US for all the mitigation scenarios. The four subplots refer to VHI and SH compliance levels of 60%, 70%, 80% and 90%. Each solid line in each subplot corresponds to a different stay-home duration. The shaded area around the solid line shows the stochasticity in the simulation results and is marked by one standard error band. The following observations can be made from figure 2: (i) a higher SH duration lowers the peak of the epidemic curve; (ii) in all cases, a longer SH duration either delays the peak and/or flattens it; (iii) a large second wave hits in Fall 2020 unless VHI and SH compliance rates are at least 80% and SH duration is at least 45 days. Note that by Fall all interventions end including schools closures; and (iv) if VHI and SH compliance rates are 90% and SH duration is 45 days or longer, the epidemic ends by the end of the year.

**Figure 2.**
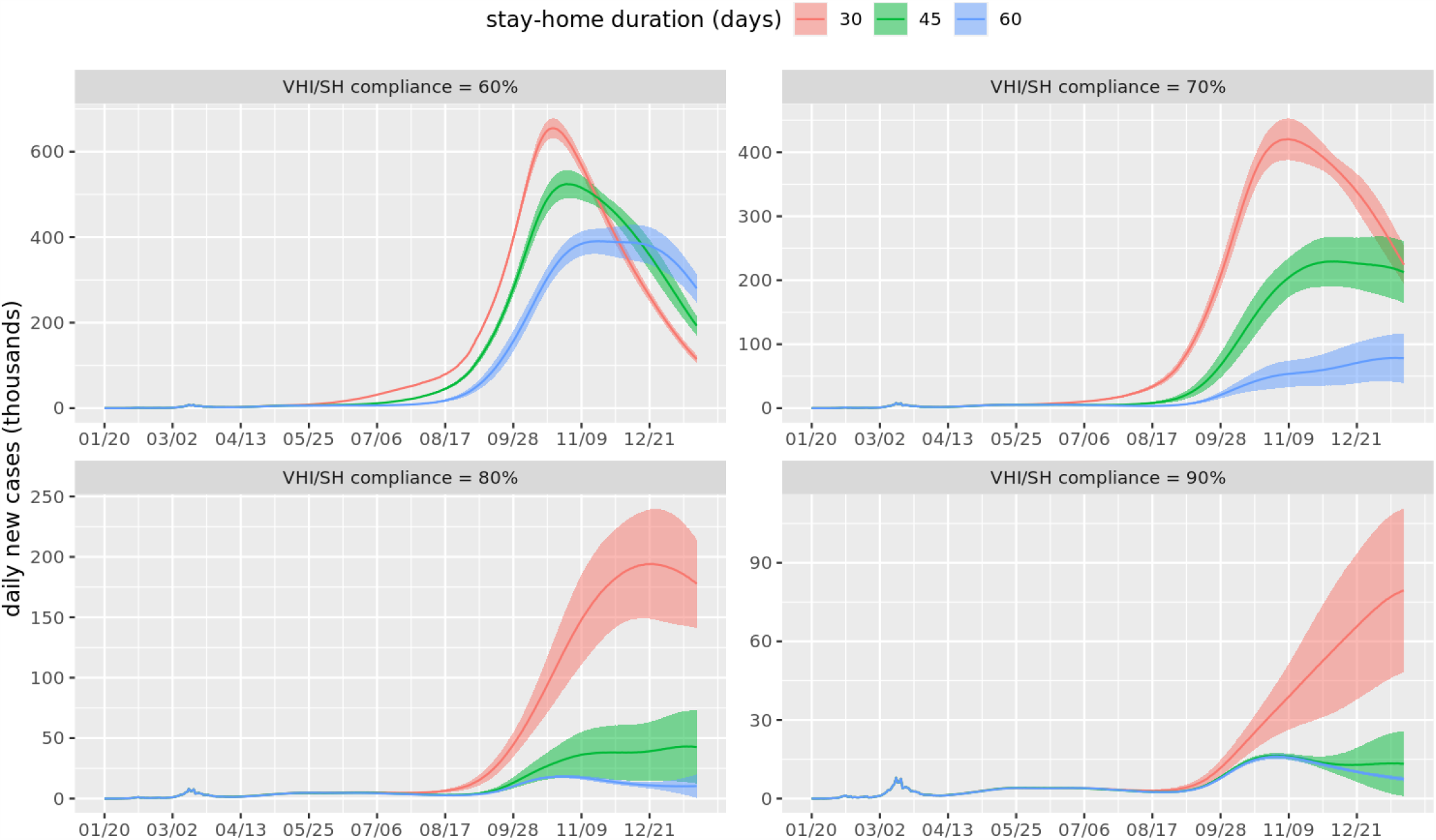
Epidemic curves showing daily new cases for the entire US, beginning in January 2020 until December 2020. Each subplot corresponds to a different value of VHI (voluntary home isolation) and SH (stay-home) compliance. Different solid lines refer to different durations of stay-home order. The shaded area around each curve is the standard error band reflecting the stochasticity in the simulation.

### 3.1 Medical cost under mitigation: Unlimited supply of hospital beds

Figure 3 shows medical costs for the 12 mitigation scenarios as well as the unmitigated one. Counts for health states, for each scenario, are estimated by our model and the simulations. Costs for categories ventilated, hospitalized and medically-attended are calculated by multiplying the per capita costs for each of these health states given in table 1 by the counts of individuals who reach that health state before recovering or dying. If a person dies, then an additional cost of death is incurred.

**Figure 3.**
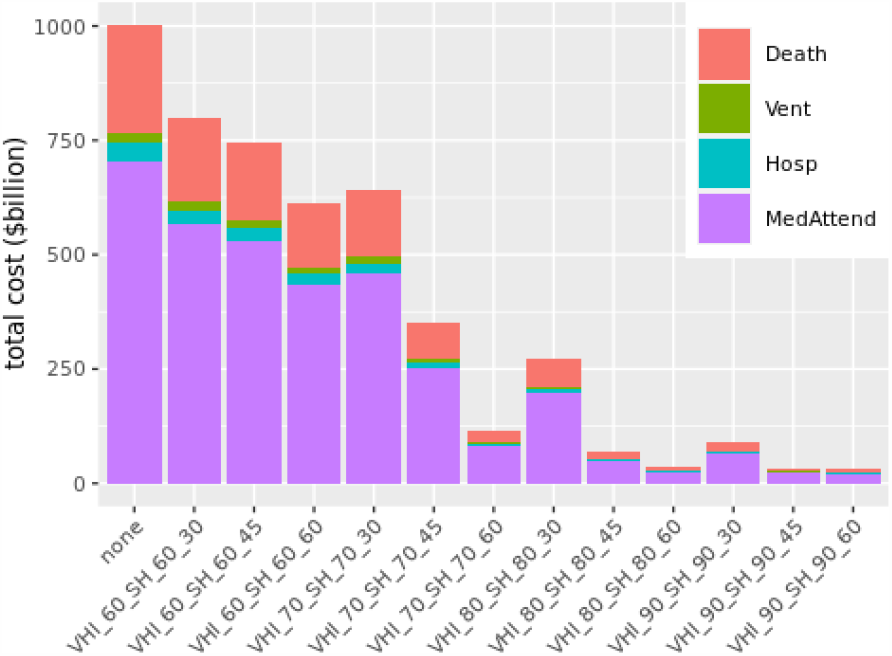
Medical costs under different mitigation scenarios and unlimited bed capacity. Here VHI refers to “voluntary home isolation”, SH refers to “stay-home”, and none refers to the unmitigated scenario. The numbers next to VHI and SH refer to their respective compliance levels and the last number represents the duration of the “stay-home” order. For example, *VHI*_60_*SH*_60_30 implies VHI compliance at 60%, SH compliance at 60% and SH duration is 30 days.

### 3.2 Medical costs: Limited availability of hospital beds and no sharing of beds allowed

Figure 4 shows medical costs that are likely to result under different mitigation scenarios given the assumption that hospital-bed capacity is limited in each HRR and is fixed at a level given by AHA. However no sharing of hospital beds is allowed outside the HRR if there is a shortage of beds. It further assumes hospital stay on average is for 14 days and 70% of the beds are dedicated to COVID-19 patients^27^. If the demand for beds exceeds the supply, patients who do not find a bed, will die. Each additional death results in extra costs at the rate of $2 millon per person. Figure 5 shows the additional deaths that are likely to be caused due to limited bed capacity. We show detailed results for the most likely scenario of 14 days of hospital stay^27^ and 70% of the bed capacity being available to COVID-19 hospitalizations. However, sensitivity analysis has been done by varying length of hospital stay to 7 days and dedicated beds availability to 80%, 90% and 120%.

**Figure 4.**
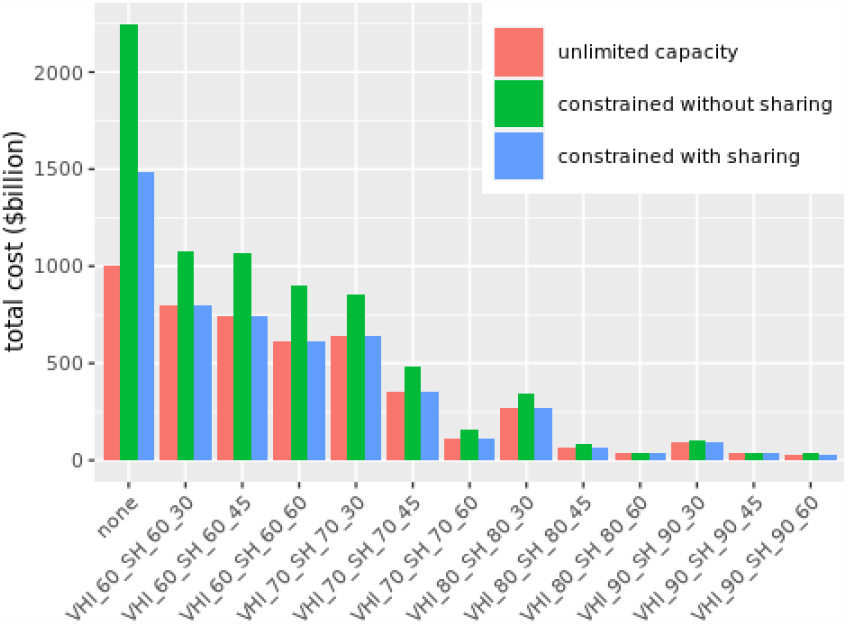
Total medical costs under different mitigation scenarios with limited and unlimited hospital bed capacity. “Constrained without sharing” refers to limited bed capacity and no sharing allowed. “Constrained with sharing” refers to limited bed capacity with sharing of beds among neighboring HRRs allowed. The constrained cases assume an average of 14 days of hospital stay and 70% of dedicated bed capacity to COVID-19 patients. X-labels are similar to those in Figure 3.

**Figure 5.**
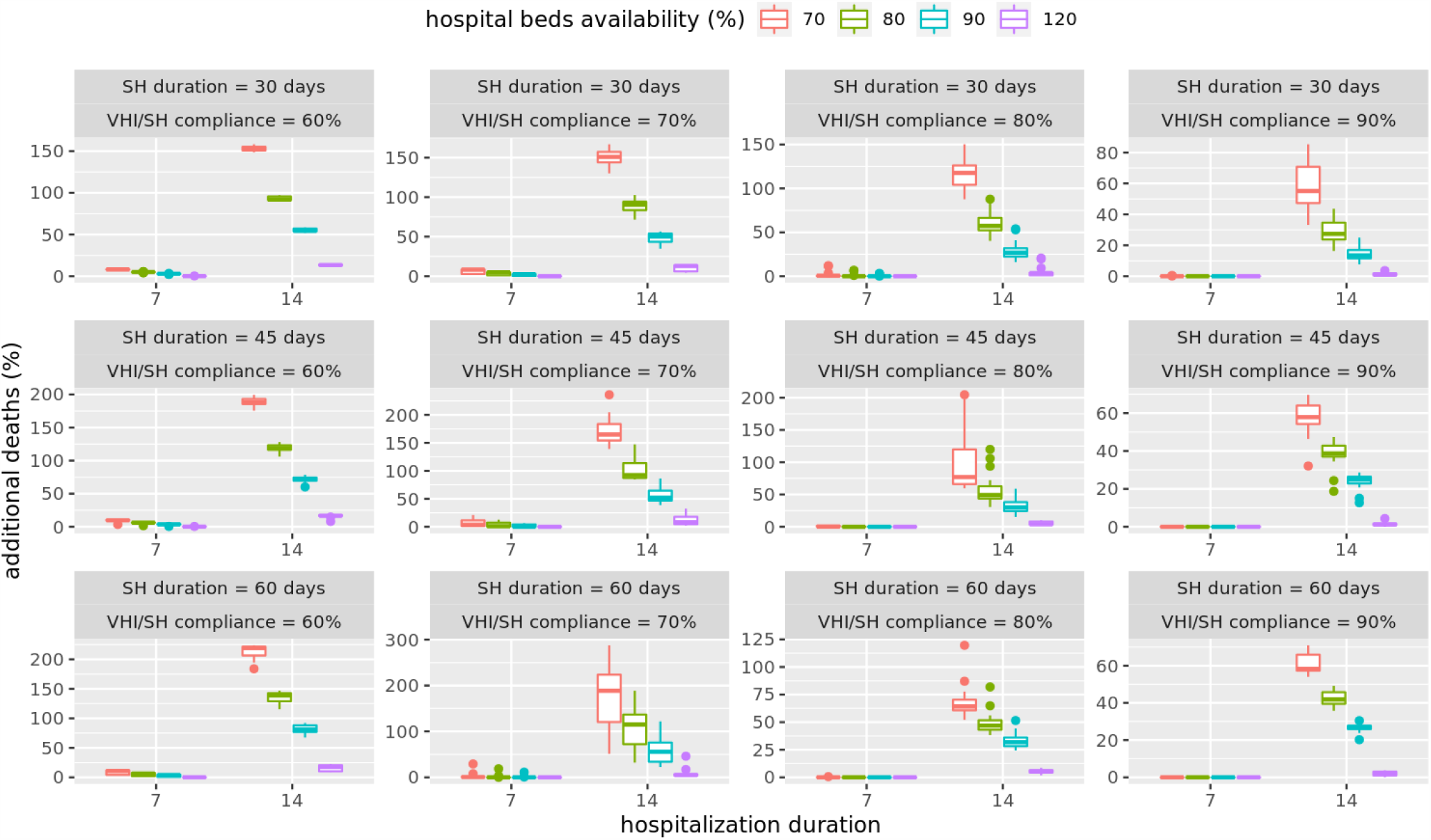
Additional deaths (compared to unmitigated scenario) under different mitigation scenarios with limited hospital bed capacity and no sharing of beds among HRRs allowed. The box plots show results for average hospital durations of 7 days or 14 days, and dedicated bed capacity of 70%, 80%, 90% or 120%.

### 3.3 Medical costs: Limited availability of hospital beds with sharing of beds allowed across regions

An analysis of costs under the assumption that hospital beds can be shared with neighboring HRRs shows that the shortage of beds can be completely alleviated by sharing. In almost all mitigation scenarios there are no additional deaths due to shortage of beds and hence no additional medical costs, as shown in figures 4 and 6. This implies that there are enough beds available locally and the deficit can be fully mitigated by sharing of beds, at least for the scenarios considered.

**Figure 6.**
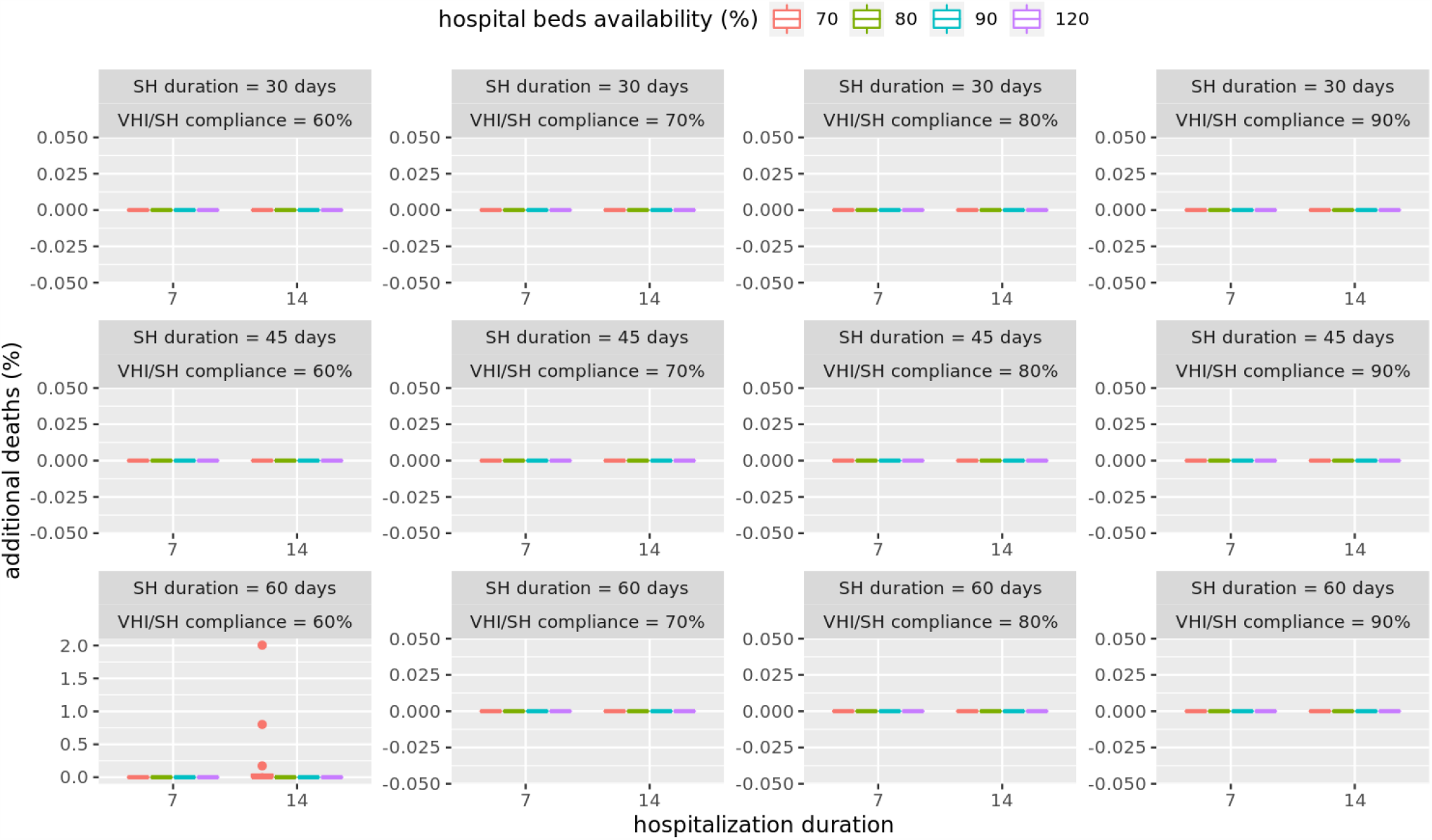
Additional deaths under different mitigation scenarios with limited hospital bed capacity and sharing of beds among neighboring HRRs allowed. The box plots shows that there are no additional deaths for average hospital durations of 7 days and 14 days, and dedicated bed capacity of 70%, 80%, 90% and 120%.

### 3.4 Sensitivity Analysis

We analyze the sensitivity of our results to multiple factors i.e. the reproductive number (*R*_0_, i.e. the expected number of cases one infected person generates), compliance to stay-home order, compliance to voluntary home isolation when symptomatic, and the duration of the stay-home order. *R*_0_ varies from 2.1 to 2.8 in increments of 0.1; VHI compliance and SH compliance vary from 30% to 90% in increments of 10% and duration of SH order varies from 30 to 75 days in increments of 15 days. Together these parameter values would generate a 1568 cell factorial experiment. To keep the analysis more manageable we use a Latin Hypercube Sampling method to generate a random sample of parameter values from this multi-dimensional distribution^28,29^, to do the sensitivity analysis. We randomly select 30 cells with different combinations of *R*_0_, VHI compliance, SH compliance and SH durations and for each cell, run 25 replicates. Medical costs are then calculated for each of the replicates for each of the 30 cells based on the health outcomes of the individuals and their averages are reported in figure 7.

**Figure 7.**
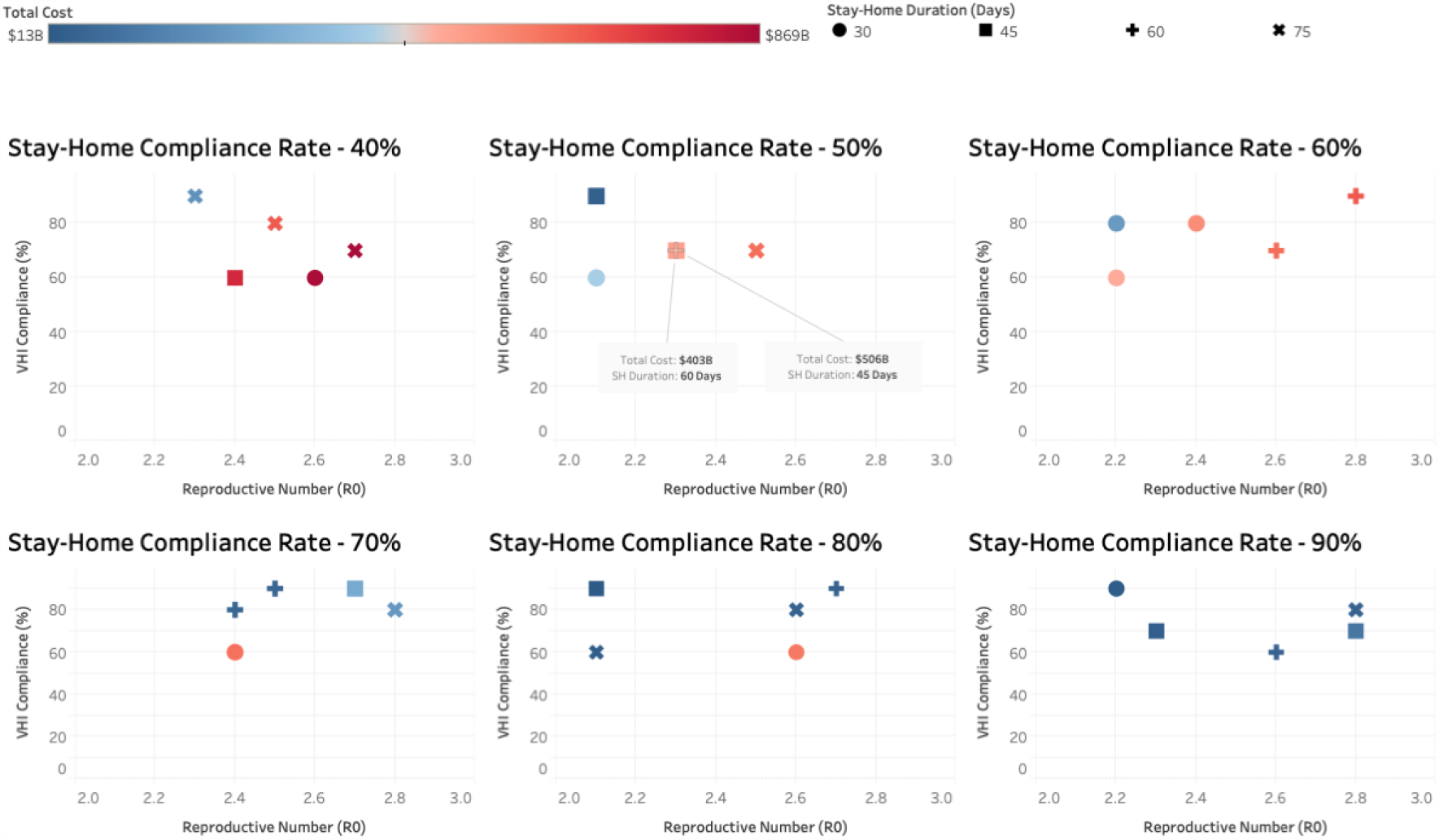
Sensitivity of total cost (in billions of dollars) to different parameter values of the reproductive number, compliance to voluntary home isolation of the symptomatic cases, compliance to stay home order, and the length of stay-home order.

The six panels in figure 7 refer to six different stay-home compliance rates of 40%, 50%, 60%, 70%, 80% and 90%. The “type” of the point refers to SH duration of 30, 45, 60 and 75 days and the color refers to the total cost. X and Y axes show *R*_0_ and VHI compliance, respectively. Results in figure 7 show that the medical costs are most sensitive to SH duration and the sensitivity of costs to *R*_0_ decreases as the SH duration increases. More discussion on sensitivity is provided in section 4.

## 4 Discussion

This analysis focuses on estimating medical costs of COVID-19 pandemic in the US under different mitigation scenarios. The results in figure 3 show that over 96% of the total medical costs could be eliminated if 90% of the people complied with the stay-home (SH) orders for 45 days or more, and followed voluntary-home-isolation (VHI) for 14 days if symptomatic, and the schools were kept closed. The unmitigated medical costs of the pandemic is over one trillion dollars i.e. about 5% of the US economy and could be brought down to just $35 billion. Similar results could be obtained if 80% of the individuals complied with SH for 60 days and followed VHI. However if compliance levels dropped to 60%, then even with a 60 day duration of SH order, total costs would still be $636 billion. To mitigate costs by at least 50%, a compliance level of at least 70% would be needed for a 45 day long stay-home order, along with VHI and SC. Figure 3 shows the trade-offs between costs, compliance levels and additional length of stay-home order under different mitigation strategies. It shows the extent to which gains could be made by an additional two weeks of lockdown and higher compliance levels. Our calculations show that the rankings of these strategies do not change if the value of life is increased from $2 million to $10 million, indicating robust rankings of scenarios. Note that the analysis in figure 3 assumes an unlimited supply of hospital beds. However if there is a limited supply of beds available as given by each HRR capacity, and no sharing of beds is allowed between HRR hospitals, then the shortage of beds can cause deaths to go up by 200% as shown in figure 5. This kind of analysis can help determine the number of temporary hospitals or hospital beds that should be arranged in advance to offset the shortage of beds.

A sensitivity analysis of the length of hospital stay, and dedicated bed capacity available to COVID-19 patients, shows that the number of deaths depend significantly on these parameters. In almost all cases, figure 5 shows 70% of dedicated bed capacity and an average 14 days hospital stay, can still lead to 100-200% increase in deaths, if the compliance is below 90% and no sharing of hospital beds is allowed. Even for the 7 days average hospital stay case, at least 70% or more compliance is needed to minimize the effect of shortage of beds on additional deaths.

Figure 6 shows results for the case when sharing of beds is allowed among hospitals in the neighboring HRR. Sharing helps eliminate almost all shortages in all mitigation scenarios. Even a 14 days hospital stay with just 70% beds available can be enough to handle the demand if sharing among HRRs is allowed. This establishes the importance of sharing medical resources during the pandemic.

In figure 7, we analyze sensitivity of medical costs to different values of *R*_0_, SH duration, VHI compliance, and SH compliance. We make several observations: (i) costs are most sensitive to SH duration. This can be seen by comparing last two panels with SH compliance rate of 80% and 90%. When *R*_0_ is 2.6 and VHI compliance rate is 60%, the costs are much higher for a 30 days SH duration (pink circle) compared to 60 days SH duration (blue +). Although SH compliance is also slightly higher for this element in the last panel (80% vs. 90%), the costs are 15 times as much for SH duration of 30 days ($597b) compared to 60 days ($39b); (ii) sensitivity of costs to *R*_0_ decreases as the SH duration increases. This is due to the fact that lockdown cuts off social contacts and reduces the effective *R*_0_ so as the length of the lockdown increases, role of *R*_0_ decreases; (iii) costs are not as sensitive to VHI compliance when SH duration is high. This is expected since social connectivity is already low when SH duration is high so low VHI compliance does not have as much of an impact.

## 5 Limitations

The costs are proxies based on pneumonia cases and are not real COVID-19 specific costs. The costs are average costs per capita and do not change by age group. For older individuals the cases are more likely to be severe and given that the distribution of infected is more biased towards older individuals, our costs only provide a lower bound on the total medical costs that are likely to occur. This research only estimates the medical costs of COVID-19 and not the costs that arise from loss in productivity from staying home, disruptions in supply chain due to lockdown, loss in travel, tourism, and hospitality industries as well as other economic activities.

## 6 Conclusions

This study estimates the medical costs of COVID-19 in the US under different mitigation scenarios and helps understand the tradeoffs between deaths, costs, infections, compliance to social-distancing and the duration of stay-home order. Our scenario-based analysis estimates the medical burden of the disease in monetary terms, and helps rank-order mitigation strategies. We show that sharing of hospital beds among neighboring “Hospital referral region” during a demand surge can reduce the additional number of deaths to almost zero. For the case where hospital referral regions do not share the hospital beds, this kind of analysis can help decide where the temporary hospitals may need to be built to offset the deficit in demand for beds.

## Data Availability

All simulated data and data processing scripts are available upon request from the corresponding author.

## Acknowledgements

The authors would like to thank members of the Biocomplexity Institute at the University of Virginia (UVA) for useful discussion and suggestions. We also thank the staff members at the UVA’s high performance computing center and at the Pittsburgh Supercomputing center for providing the much needed high-performance computing resources. This work was partially supported by National Institutes of Health (NIH) Grant R01GM109718, NSF BIG DATA Grant IIS-1633028, NSF DIBBS Grant ACI-1443054, NSF Grant No.: OAC-1916805, NSF Expeditions in Computing Grant CCF-1918656, CCF-1917819, US Centers for Disease Control and Prevention 75D30119C05935, DTRA subcontract/ARA S-D00189-15-TO-01-UVA, and a collaborative seed grant from the UVA Global Infectious Disease Institute. The content is solely the responsibility of the authors and does not necessarily represent the official views of the sponsoring agencies.

## Contributions

JC, SH, HM, SE, SV and BL built the model and the software. AV, JC, WY and AM processed and analyzed the data. AV, JC, AM, MM and CB conceived the project. All authors helped write, edit and review the paper.

## Ethics declarations

The authors declare no competing interests.

## Appendix: The Disease Model Parameters

The CDC disease model in Figure 1 shows the state transitions which include transmissions and progressions. The former occurs when an individual in *Susceptible* state comes in contact with an individual in one of *Presymptomatic, Symptomatic*, or *Asymptomatic* states. The latter occurs when an individual has been in that state for a certain amount of time (called dwell time); the transitions are probabilistic. The dwell time distributions and the transition probability distributions are age dependent and are specified in Table 4.

**Table 4.** Age dependent dwell-time distributions and transition probability distributions. Here “dt” refers to duration, e.g. dt-mean is mean duration and dt-discre1t5e is discrete distribution duration. “Attd” is medically attended; “Hosp” is hospitalized; “Vent” is ventilated; “Sympt” is symptomatic and “Asympt” is asymptomatic.

| Progression | Attribute | Age group |  |  |  |  |
| --- | --- | --- | --- | --- | --- | --- |
|  |  | 0-4 | 5-17 | 18-49 | 50-64 | 65+ |
| Exposed - Asympt | prob |  |  | 0.35 |  |  |
| Exposed - Asympt | dt-mean |  |  | 5 |  |  |
| Exposed - Asympt | dt-std dev |  |  | 1 |  |  |
| Asympt - Recovered | prob |  |  | 1 |  |  |
| Asympt - Recovered | dt-mean |  |  | 5 |  |  |
| Asympt - Recovered | dt-std dev |  |  | 1 |  |  |
| Exposed - Presympt | prob |  |  | 0.65 |  |  |
| Exposed - Presympt | dt-fixed |  |  | 1 |  |  |
| Presympt - Sympt | prob |  |  | 0.65 |  |  |
| Presympt - Sympt | dt-fixed |  |  | 1 |  |  |
| Sympt - Attd | prob | 0.9594 | 0.9894 | 0.9594 | 0.912 | 0.788 |
| Sympt - Attd | dt-discrete | 1:0.175, 2:0.175, 3:0.1, 4:0.1, 5:0.1, 6:0.1, 7:0.1, 8:0.05, 9:0.05, 10:0.05 |  |  |  |  |
| Attd - Recovered | prob |  |  | 1 |  |  |
| Attd - Recovered | dt-mean |  |  | 5 |  |  |
| Attd - Recovered | dt-std dev |  |  | 1 |  |  |
| Sympt - Attd(D) | prob | 0.0006 | 0.0006 | 0.0006 | 0.003 | 0.017 |
| Sympt - Attd(D) | dt-fixed |  |  | 2 |  |  |
| Attd(D) - Hosp(D) | prob |  |  | 0.95 |  |  |
| Attd(D) - Hosp(D) | dt-fixed |  |  | 2 |  |  |
| Hosp(D) - Vent(D) | prob | 0.06 | 0.06 | 0.06 | 0.15 | 0.225 |
| Hosp(D) - Vent(D) | dt-fixed |  |  | 2 |  |  |
| Vent(D) - Death | prob |  |  | 1 |  |  |
| Vent(D) - Death | dt-fixed |  |  | 4 |  |  |
| Hosp(D) - Death | prob | 0.94 | 0.94 | 0.94 | 0.85 | 0.775 |
| Hosp(D) - Death | dt-fixed |  |  | 6 |  |  |
| Attd(D) - Death | prob |  |  | 0.05 |  |  |
| Attd(D) - Death | dt-fixed |  |  | 8 |  |  |
| Sympt - Attd(H) | prob | 0.04 | 0.01 | 0.04 | 0.085 | 0.195 |
| Sympt - Attd(H) | dt-fixed |  |  | 1 |  |  |
| Attd(H) - Hosp | prob |  |  | 1 |  |  |
| Attd(H) - Hosp | dt-mean | 5 | 5 | 5 | 5.3 | 4.2 |
| Attd(H) - Hosp | dt-std dev | 4.6 | 4.6 | 4.6 | 5.2 | 5.2 |
| Hosp - Recovered | prob, |  |  | 0.2 |  |  |
| Hosp - Recovered | dt-mean | 3.1 | 3.1 | 3.1 | 7.8 | 6.5 |
| Hosp - Recovered | dt-std dev | 3.7 | 3.7 | 3.7 | 6.3 | 4.9 |
| Hosp - Vent | prob | 0.06 | 0.06 | 0.06 | 0.15 | 0.225 |
| Hosp - Vent | dt-mean |  |  | 1 |  |  |
| Hosp - Vent | dt-std dev |  |  | 0.2 |  |  |
| Vent - Recovered | prob |  |  | 1 |  |  |
| Vent - Recovered | dt-mean | 2.1 | 2.1 | 2.1 | 6.8 | 5.5 |
| Vent - Recovered | dt-std dev | 3.7 | 3.7 | 3.7 | 6.3 | 4.9 |

## Notes

### Competing Interest Statement

The authors have declared no competing interest.

### Author Declarations

The work does not include any human subjects.

